# Covid Fast Fax: A system for real-time triage of Covid-19 case report faxes

**DOI:** 10.1101/2020.12.15.20248256

**Authors:** Adam Lavertu, Alison Stribling, Matt White, Greg McInnes, Russ B. Altman, Rajiv Pramanik, Amit Kaushal

**Affiliations:** Stanford University, Department of Biomedical Data Science, Stanford CA; Contra Costa Health Services, Martinez, CA; Stanford University, Department of Bioengineering, Stanford, CA

## Abstract

The scale and speed of the COVID-19 pandemic has strained many parts of the national healthcare infrastructure, including communicable disease monitoring and prevention. Many local health departments now receive hundreds or thousands of COVID-19 case reports a day. Many arrive via faxed handwritten forms, often intermingled with other faxes sent to a general fax line, making it difficult to rapidly identify the highest priority cases for outreach and monitoring. We present an AI-based system capable of real-time identification and triage of handwritten faxed COVID-19 forms. The system relies on two models: one model to identify which received pages correspond to case report forms, and a second model to extract information from the set of identified case reports. We evaluated the system on a set of 1,224 faxes received by a local health department over a two-week period. For the 88% of faxes of sufficient quality, the system detects COVID-19 reports with high precision, 0.98, and high recall, 0.91. Among all received COVID-19 faxes, the system identifies high priority cases with a specificity of 0.87, a precision of 0.46 and recall of 0.83. Our system can be adapted to new forms, after a brief training period. Covid Fast Fax can support local health departments in their efforts to control the spread of COVID-19 and limit its impact on the community. The tool is freely available.

## Introduction

In the United States, local health departments (LHDs) are the agencies responsible for managing the public health response to COVID-19 cases in communities across the country. Once a treating provider or a laboratory notifies the LHD of a positive COVID-19 individual, the LHD must carry out several functions related to the notified case. These include case outreach, identification of gaps in care, assistance with isolation, contact tracing, contact outreach and quarantining, and others.

Some case reports necessitate an urgent or emergent response. For example, a report of an outbreak in a congregate living setting with a vulnerable population, such as a skilled nursing facility^1^ or homeless shelter^2^, may necessitate an immediate mobilization of resources to identify potentially exposed individuals, facilitate isolation of cases, and testing and quarantining of potentially exposed individuals.

There are at least two factors that make it difficult for LHDs to quickly identify and triage high-acuity COVID-19 cases. The first is the sheer volume of COVID-19 cases. As of this writing, there are over 15 million confirmed COVID-19 cases in the United States, with new cases accruing at over 1 million per week^3^. Public health department staffing and systems have historically not been resourced to deal with a disease spreading at this scale or speed.

The second is the lack of a universal electronic case reporting and triaging system^4^. Throughout the country, a large portion of COVID-19 cases are reported to LHDs via case reporting forms known as confidential morbidity reports (CMR). These CMR forms are often filled out by hand and faxed. Multiple different CMR form versions may be used to report cases to the LHD. In contrast to an electronic reporting interface, which would enable health officials to sort and filter cases matching specific criteria to guide acuity, each handwritten fax must be manually reviewed. These faxes are usually processed in the order received; therefore, a report of a high-acuity case may sit in the queue of incoming faxes for hours or days before it is processed. Many LHDs now have trained public health officials monitoring and reading incoming faxes throughout the day, a costly use of limited public health resources^5^.

Advances in the computational fields of artificial intelligence (AI) and machine learning (ML) have enabled the design of computer algorithms that can effectively classify images and identify handwritten characters and digits^6,7^. In this work, we demonstrate an AI-based system and associated alerting workflow which scans all inbound faxes received by an LHD fax line, detects the subset of faxes corresponding to COVID-19 reporting forms, “reads” key elements of the form to determine level of acuity based on criteria set by public health officials, and can initiate an automated email alert to LHD officials when a high-acuity fax is received, in real-time (Figure 1). We describe technical performance of our case reporting form detection algorithm and acuity detection algorithms, and describe initial results from operational deployment.

**Figure 1:**
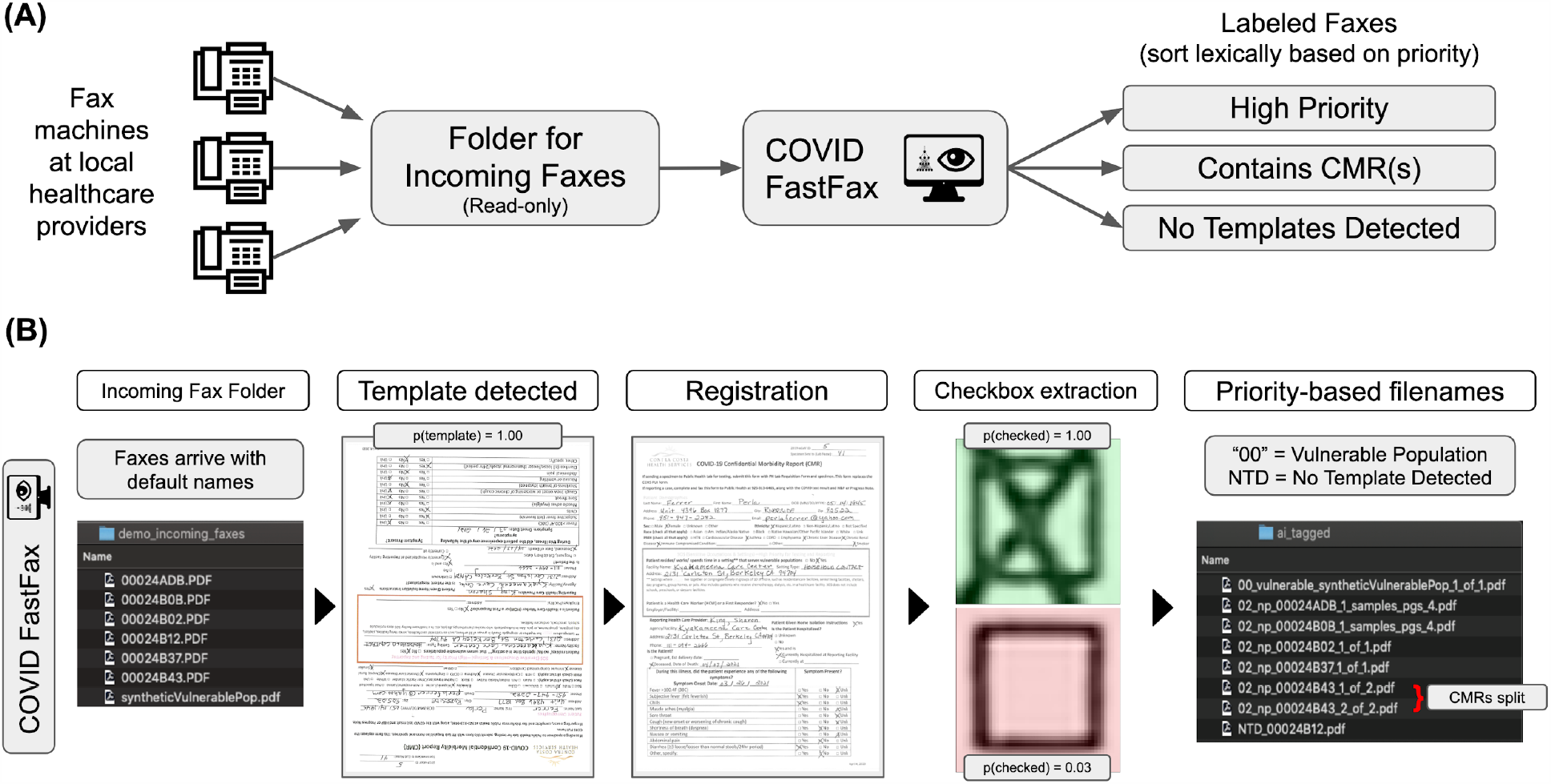
Overview of COVID Fast Fax System. (A) Shows overall CMR fax workflow, where local healthcare providers faxes are received by a holding folder that maintains the original faxes unaltered. These fax documents are then ingested by the COVID Fast Fax system yielding prioritized reports based on priority status and likelihood of containing a CMR. (B) Schematic showing the COVID Fast Fax internal system process. An ingested PDF is split into pages and each page is processed separately. First, a neural network classifies each page as to whether it contains one of the CMR templates. Second, if a template is detected, the page is registered to the relevant known templates and a match is confirmed. If a match is confirmed, then checkboxes are extracted and classified as checked or unchecked. This process results in metadata on triage priority, the total number of CMRs, and the location of CMRs within the original documents.

## Methods

A local LHD fax line was selected for study. Faxes are received digitally and stored as PDFs on a network folder. The fax line received CMR forms, sometimes individually and other times in batches, as well as other fax documents.

### CMR form identification model

In order to extract information from CMR forms, we first developed a model to recognize which pages from the received fax line were CMR forms and which were other documents. We aimed to train this CMR form identification model for the most frequently used CMR forms. To identify these forms, LHD staff intermittently sampled incoming fax reports from the incoming fax line for a period of 2 days. Some form templates were used exclusively by providers that switched from fax to digital reporting during the period of the study; these forms were excluded. The 5 most frequently used forms were identified.

A desired feature of the training pipeline was training models without using actual completed CMR forms, owing to the sensitive nature of the PHI contained in the reports. Therefore, only a single unfilled example of each of the 5 CMR form templates, in pdf format, was used to initiate algorithm development.

To create a training set, 5,000 copies of each CMR template were created. Each of these images was normalized then heavily augmented in an attempt to mimic various forms of noise observed from fax systems (blur, noise, pixel dropout, random cutouts, affine transformations, streaks). A negative set was generated by sampling a set of 5,000 PDF images (e.g. textbook pages, scientific papers, etc.) from the downloads folder of one of the authors (AL).

We trained a neural network to perform CMR identification and classification. We used PyTorch 1.4.0 to construct and train several small convolutional neural networks^8,9^. The specific model architecture can be found in the GitHub repository for this project, within the TemplateNet class in `models.py`. The two networks that performed best on classifying a held-out set of negatives vs augmented template images were used in the final system. We use the union of their non-negative predictions as the set of predicted templates for a page of the PDF in the final system.

### CMR Data Extraction Model

#### Synthetic report data collection and preprocessing

We selected 2 of the 5 templates to use for generating data to train our CMR data extraction models. To generate training examples of handwritten and completed forms, volunteers filled out forms each with simulated case data, which was provided to them, and faxed the result back to a fax line we used for research purposes.

To generate simulated case data we wrote a Python program to generate synthetic data for every field of the CMR form. We used the Faker package (https://github.com/joke2k/faker) to simulate names and contact information. Hospitals and their associated data were sampled from a list of hospitals in California. Date fields were sampled from March 1st, 2020 forward and required that dates of events logically correspond to the sequence of events (e.g. first symptoms sometime in two-weeks prior to hospital visit, patient death cannot occur prior to hospitalization, etc.). Checkbox fields were sampled randomly from the available options on each form, attempting to be logically consistent (i.e. checking “Ever in ICU”, required that “Ever hospitalized” also be checked).

13 volunteers completed 10 simulated cases each, for a total of 130 completed CMR forms. No real PHI was used to generate these 130 handwritten training examples.

We mapped coordinates of pixels in each form template to their respective fields and checkboxes. We extracted the relevant regions of each of the collected sample forms for training the checkbox models with a buffer of 10 pixels from the center of the template.

#### Checkbox model training framework

Much of the actionable information in the CMR forms is reported in structured data fields, often with checkboxes. For example, check boxes denote whether a case resides in a congregate living facility or is a member of a vulnerable population. We built a model to identify which checkboxes on a form are checked.

We sorted checkboxes into checked (positive) and unchecked (negative) classes based on the ground truth synthetic data. We split the checkbox data into training and validation samples at the writer level, with no overlap of writers between the training and validation sets. We normalized the checkbox images and augmented them during training with random flips, gaussian blur, and gaussian noise. We trained five small convolutional neural networks for 50 epochs each and kept the models with the best validation loss. The specific model architecture can be found in the GitHub repository for this project, within the CheckNet class in `models.py`. We combined these fives models into an ensemble classifier. The goal of the checkbox model is to identify cases that are high priority for follow-up. False positives are of little consequence as all CMRs are eventually processed, but false negatives may not be processed for hours or days, potentially resulting in additional spread in vulnerable populations. Thus, we placed our emphasis on sensitivity over specificity, to avoid false negatives, and classified any sample where at least two of the five models in the ensemble predicted a check as checked.

### CMR high acuity case definition

LHD officials identified a set of criteria that would designate a case high acuity. Criteria included working or living in a setting that serves populations vulnerable to severe COVID-19 and/or being a healthcare worker.

### Overall system implementation

We implemented the entire system in Python 3.6.11. The user specifies a directory to monitor and the system automatically ingests new PDFs that appear in that directory in a read-only manner. The template classification model makes a prediction for each page of the PDF.

Pages that are predicted to be a relevant CMR are then registered to the predicted form template using the `pyStackReg` (https://github.com/glichtner/pystackreg) and `imreg_dft` (https://github.com/matejak/imreg_dft) Python packages. We found precise template registration to be a consistent issue and settled on multiple registrations. For the first registration, a binarized image is first registered to the template using only scale and rotation transformations, then a second affine transformation based registration is performed on the result of the first registration. The transformation matrices for each of these steps are stored then applied in sequence to the original PDF image, resulting in a registered non-binarized image. Registered images are then compared to the template image using normalized cross-correlation via the `match_template` function in the `skimage` library, using the center 80% of the template image to match against^10^. The L1 distance between the location of the center section in the registered image versus its location in the template image is calculated. An L1 distance of less than 100 pixels and a max `match_template` correlation coefficient of greater than 0.5 is considered a match and is processed further. If the first registration attempt fails, we run a second registration with `imreg_dft` and consider a template a match if the L1 distance is less than 100 pixels and a max `match_template` correlation coefficient of greater than 0.25.

We extract the checkbox regions from the registered image by identifying peaks in the normalized cross-correlation between a small region surrounding the desired checkbox in the template image and a slightly larger region in the same location in the registered image.

### Mirror testing

The system was deployed in a LHD virtual machine on November 2, 2020 and ran for 14 days until November 16, 2020. PDFs were sorted by a human reviewer based on whether they contained a CMR report template or not, if they contained a CMR report template they were further sorted into which of the 3 templates they contained, ignoring different subversions of each form (i.e. California CMRs for March and April were both sorted into the California CMR category). Accuracy of the system was then assessed on the ability of the system to detect a template form and the correct acuity for each CMR report. Acuity statistics were calculated using all forms, including those CMRs not detected at the template detection stage.

To rapidly inform county health officials of high-acuity cases, an email alert was sent whenever a high-acuity case was identified. During testing, this email was sent initially to one of the coauthors (AL) for manual confirmation, then forwarded on to the appropriate LHD officials.

#### IRB exemption

This project was deemed exempt by the Stanford IRB.

## Results

We present a system for the real-time triaging of Covid case reports received via fax (Figure 1). The system is capable of handling five different report templates. The five templates were selected based on a sample of 186 reports from incoming faxes over 2 days, from which the LHD staff identified the most frequently used CMR forms. Some of the form templates were used exclusively by providers that switched from fax to digital reporting during the period of the study (109/186); these forms were excluded. The five next most abundant forms collectively accounted for 97.4% of all eligible CMR forms sent (Figure 2).

**Figure 2:**
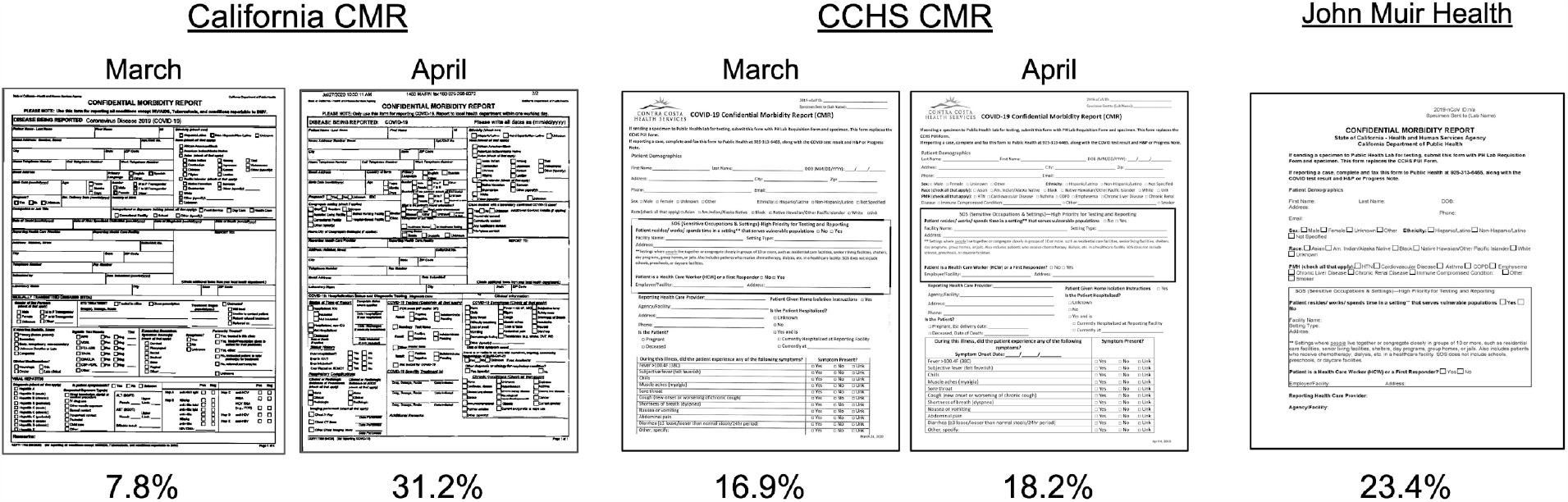
Top 5 most abundant faxed forms received over 2 day observation period. The California State CMR form, March (7.8%) and April (31.2%) versions, the CCHS CMR, March (16.9%) and April (18.2%), and the reporting format used by John Muir Health (23.4%).

We present results of our system’s performance on 1,224 faxes received in real-time via the LHD fax line over a period of 14 days. Overall, the system achieved good performance on the CMR recognition and case acuity triage tasks. We calculated performance on the document and page level, additionally reporting document level performance broken down by template. For document classification, our system detected a relevant CMR with precision of 0.98 and recall of (Figure 3A). A significant portion (142 reports, 74%) of the false negatives for CMR recognition task were extremely low-quality faxes (Supplementary Figure 1). Excluding those faxes, and limiting our performance assessment to the 88% of faxes of sufficient quality, our system detected a relevant CMR with precision of 0.98 and recall of 0.90 (Figure 3D). We calculated the page level performance statistics for our system based on the total number of pages contained within all of the 1,224 received faxes. For page level performance, our system detected a relevant CMR page with precision of 0.98 and recall of 0.70 (Figure 3B), and with precision of 0.98 and recall of 0.90 (Figure 3E), excluding the low-quality faxes. We note the large number of pages that were processed relative to the number of documents received, with each document containing an average of ∼2.80 pages. We achieved similar performance on all the templates, with precision and recall of 0.98, 0.67, for the California Statewide CMR, 1.0, for the CCHS CMR, and 1.0, 1.0 for the JMH template (Figure 3C). Only the California Statewide CMR was impacted by the low-quality fax issue, and removing those faxes we achieved precision and recall of 0.98, 0.91 averaging over all templates.

**Figure 3:**
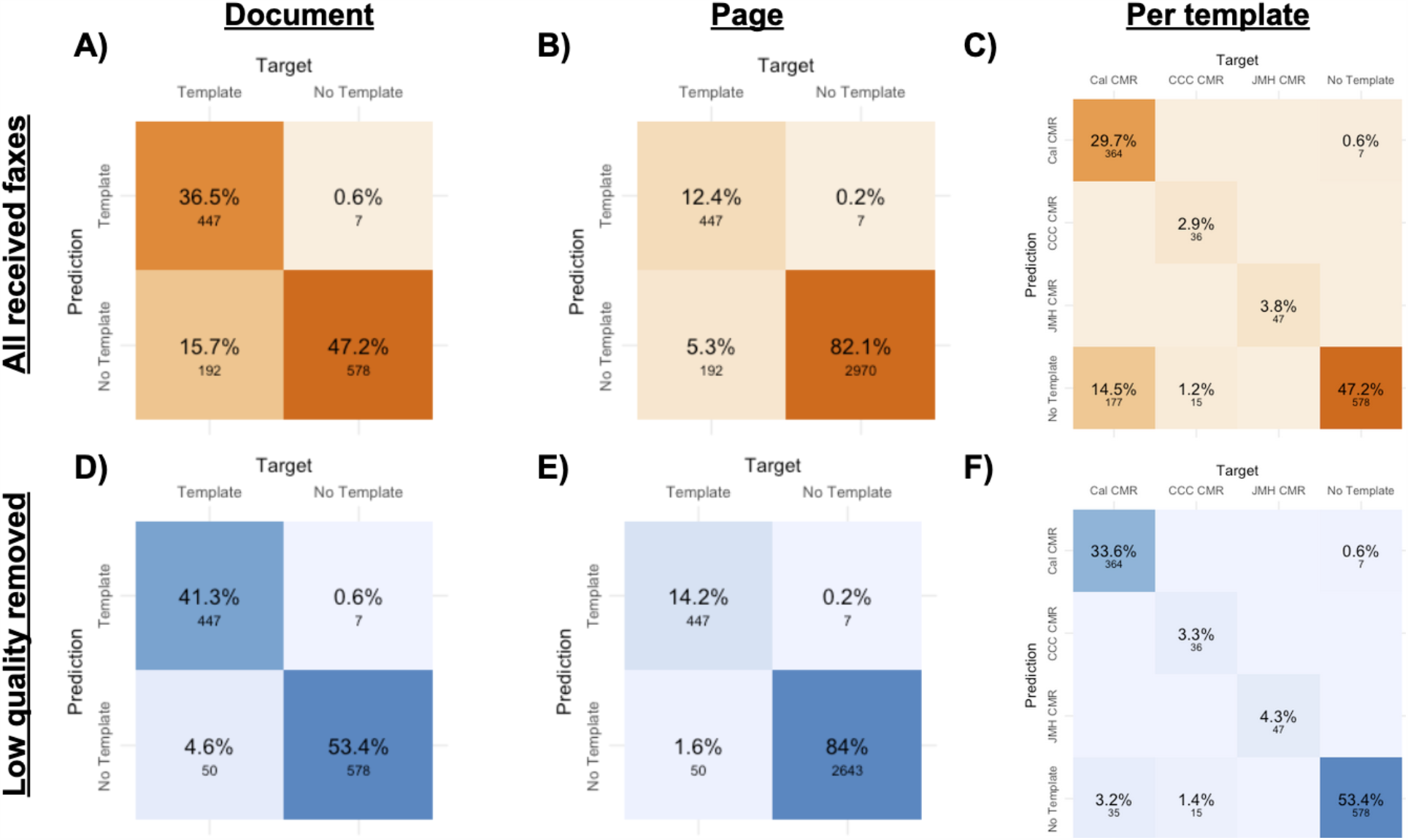
Classification performance on 1,224 faxed reports, page level and document level collected from live fax folder. Performance is shown for neural network template classifier with registration and match confirmation based filtering. (A) Document level performance across all templates. (B) Page level performance across all templates. (C) Document level performance for each template. (D) Document level performance across all templates without low-quality faxes (E) Page level performance across all templates without low-quality faxes (F) Document level performance for each template without low-quality faxes.

We assessed performance of our system on the CMR case acuity task based on its ability to accurately triage high acuity cases from the 497 CMRs received, even if those CMRs were not accurately detected by the system (Figure 4). The inclusion of CMRs that failed at the template detection stage, lowers overall performance as there is no way for the system to triage a case that was not recognized. On the CMR triaging task, we achieved good specificity of 0.87, modest precision of 0.46 and good recall of 0.83.

**Figure 4:**
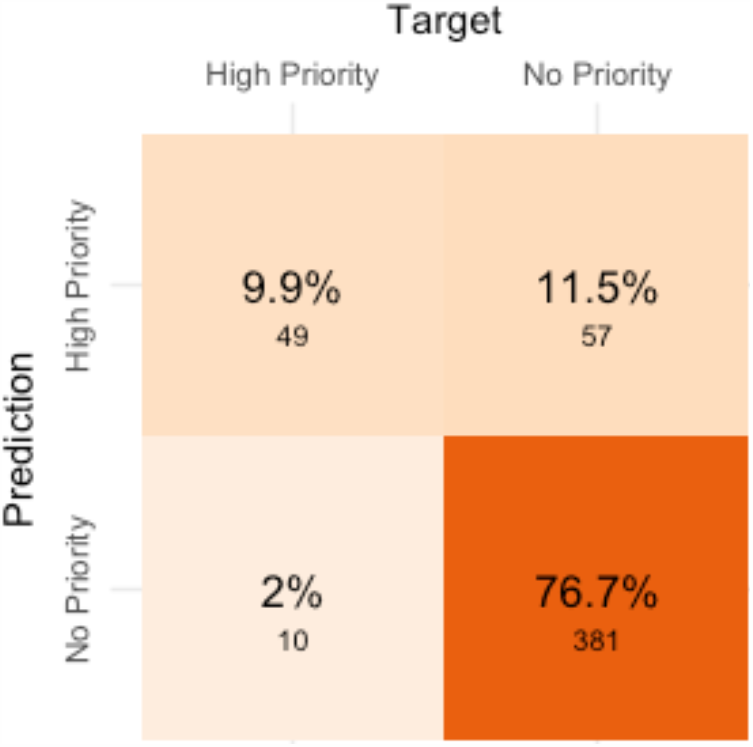
Confusion matrix of performance on prioritization of 497 faxed Covid CMR reports of sufficient quality collected from a live fax folder. Confusion matrix showing checkbox classification performance of checkbox extraction and neural network classification process.

## Discussion

Of the 3,616 pages contained in the 1,224 documents received, our algorithms were able to capture 49 of the 59 high acuity reports, with limited false positives. Reports deemed to be high acuity are moved to the front of the processing queue; by identifying the true positives quickly and reprioritizing them to the top of the queue, we hope to reduce time to identification of high acuity cases, thereby enabling a faster and more effective public health response. This approach of using AI-based classification to reorder a workflow to reduce delays in processing high acuity cases has been successfully used in other domains, for example, radiology.^11^

Our system benefits from the fortunate fact that, for the forms used, the information used to determine the acuity of a case is contained in structured fields (checkboxes). Forms that captured this same information in free-text fields instead of structured fields would pose a much more challenging problem. The ability to extract free-text information on these forms could enable further efficiency, especially related to saving time for data entry, and is a potential avenue for future work.

The checkbox detection models were developed using only the synthetic data. We have already successfully generalized the models to a new form template which was not included in the training process.

We developed this system with a limited number of training examples, using only the 5 template forms with heavy data augmentation, to train the template detection classifier. The template detection classification is a crucial step in the process as it would be too time expensive to perform template registration-based template matching on every page. The system performs well, recovering most forms of sufficient quality. The ability to reliably detect faxes that contain CMRs enables new cases to be dealt with prior to faxes containing lab reports, morbidity reports for less contagious diseases, and faxes containing advertisements or other non-relevant material. Ideally, this leads to a general improvement in the workflow efficiencies at the LHD.

We have initiated a pilot deployment of our system at a single LHD. While full operational performance metrics will be studied and reported separately, initial response to the system has been positive. The system went live prior to a long holiday weekend. The team had over 400 faxes to process upon return. The system alerted the team to several high-acuity cases which would not have otherwise been processed until ∼20 hours later.

To limit the need to change workflows or adopt new software, our system can reorder faxes simply by prepending short strings to filenames so that when files are lexically (alphabetically) sorted by the filesystem, the highest acuity files are naturally listed first.

As several LHDs face similar problems (and have similar workflows) when dealing with incoming faxes, our system has the potential to be useful in other departments as well. With existing infrastructure already developed, repurposing algorithms involves obtaining a copy of a new form and training the algorithm to detect the form and to know where the checkboxes are; this process should take less than 24 hours.

We recognize the system as it currently exists has several limitations. We are limited by the quality of the faxes being received and have no effective means to process low-quality faxes. This issue could be addressed through coordination between the LHD and regional healthcare providers to improve the quality of the faxes being sent. Additionally, image registration takes up a large amount of the current compute time of the system, while using a neural network to quickly identify which images should be registered provides a large improvement in system processing times, the need to retrain the neural network to identify new templates limits the ability of our system to generalize to other LHD in other states or counties out of the box. However, template detection and image registration are upstream of all other tasks and therefore it’s crucial they maintain high performance.

Additionally, in the future medical efforts where a high volume of faxes may be received and AI-based assistance in triaging is desired, the usage of a standardized form that is designed for machine reading would further improve the performance of systems like the one outlined here. Key components would be isolating key information, such as checkboxes that are tied to high priority cases, away from other parts of the form would facilitate machine-based extraction. The inclusion of optical character recognition oriented written fields could make the deployment of text extraction easier, as model performance on individual character recognition is already quite high. The inclusion of a QR code, or some computationally recognizable template tag could prevent the need for the template classification model and reduce system development times.

Future work will involve improving performance of existing models, the ability of the system to generalize to new templates, as well as developing a more full-featured optical character recognition pipeline to assist with data extraction and data entry, tasks that currently consume a significant amount of LHD staff time. We make our code, as well as the Template Classification and Checkbox classification models, V1.0 is benchmarked in this paper, publicly available here: https://github.com/alavertu/CovidFastFax

## Data Availability

Covid reports are PHI, so can not be made available.
Code related to the project is publicly available

https://github.com/alavertu/CovidFastFax

## Contributions

A.L., A.S., M.W., R.A., R.P. and A.K. conceived of the system. A.L., G.M., A.K., R.A. collected the synthetic report examples. A.L. designed, implemented, and evaluated the system. A.L., A.S., and M.W. deployed the system on the LHD servers. A.L., A.S., M.W., G.M., R.A., R.P. and A.K. helped draft and revise the manuscript.

---

**Supplementary Figure 1:**
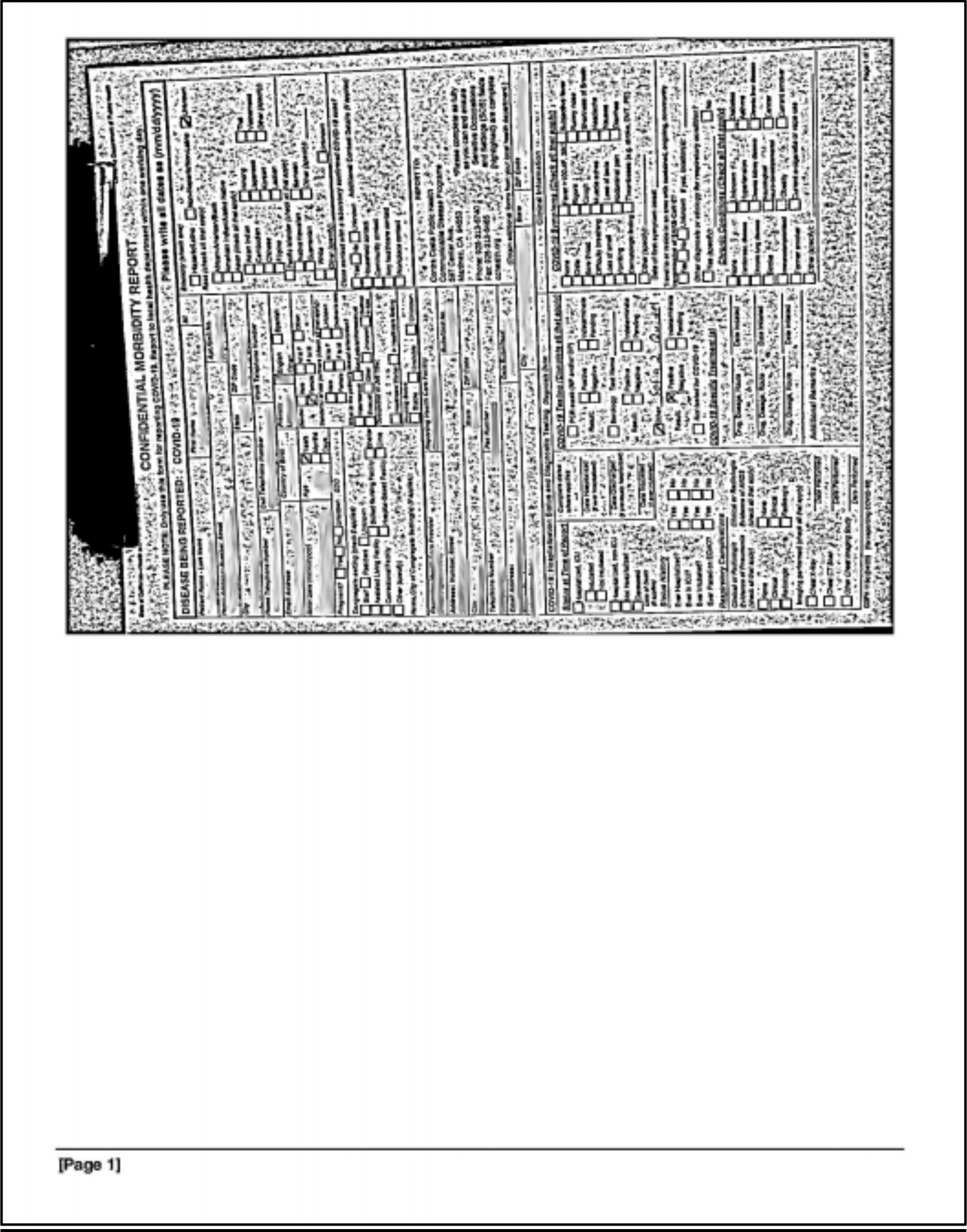
Example of a low-quality fax that the system fails to detect. A portion of the incoming faxes received over the performance evaluation phase of the deployment contained CMR forms as shown below.

## References

1. Barnett, M. L. & Grabowski, D. C. Nursing Homes Are Ground Zero for COVID-19 Pandemic. JAMA Health Forum 1, e200369–e200369 (2020).

2. Tsai, J. & Wilson, M. COVID-19: a potential public health problem for homeless populations. Lancet Public Health 5, e186–e187 (2020).

3. CDC. COVID-19 Cases, Deaths, and Trends in the US. https://covid.cdc.gov/covid-data-tracker/ (2020).

4. Rajeev, D. et al.. Development of an electronic public health case report using HL7 v2.5 to meet public health needs. J. Am. Med. Inform. Assoc. 17, 34–41 (2010).

5. Kliff, S. & Sanger-Katz, M. Bottleneck for U.S. Coronavirus Response: The Fax Machine. The New York Times (2020).

6. Lecun, Y., Bottou, L., Bengio, Y. & Haffner, P. Gradient-based learning applied to document recognition. Proc. IEEE 86, 2278–2324 (1998).

7. Deng, J., Dong, W., Socher, R., Li, L. J. & Li, K. Imagenet: A large-scale hierarchical image database. 2009 IEEE conference (2009).

8. Paszke, A. et al. PyTorch: An Imperative Style, High-Performance Deep Learning Library. in Advances in Neural Information Processing Systems (eds. Wallach, H. et al.) vol. 32 8026–8037 (Curran Associates, Inc., 2019).

9. Fukushima, K. Neocognitron: A self-organizing neural network model for a mechanism of pattern recognition unaffected by shift in position. Biological Cybernetics vol. 36 193–202 (1980).

10. van der Walt, S. et al.. scikit-image: image processing in Python. PeerJ 2, e453 (2014).

11. Annarumma, M. et al.. Automated Triaging of Adult Chest Radiographs with Deep Artificial Neural Networks. Radiology 291, 196–202 (2019).

